# Switching from algorithm-based to universal admission screening for COVID-19 in hospital settings

**DOI:** 10.1101/2020.08.07.20170001

**Authors:** Tjibbe Donker, Hajo Grundmann, Fabian Bürkin, Hartmut Hengel, Hartmut Bürkle, Thorsten Hammer, Frederik Wenz, Winfried Kern

## Abstract

Detection of COVID-19 positive cases on admission to hospitals is crucial to protect patients and staff at the same time. While universal admission screening can prevent more undetected introductions than the algorithm-based screening, which preselects patient based on their symptoms and exposure, it is a more costly strategy as it involves testing a large number of patients.

We construct a simple tool to help determine when the benefit of additionally found cases outweighs the cost of the additionally tested patients, based on the numbers of patients to be screened in an acceptable time span to find an additional case when screening all admitted patients.

## Introduction

Pre- or asymptomatic SARS-CoV-2 patients whose infections remain unrecognised at the time of hospital admission are a potential threat for patient safety and occupational health. If infectious, these patients may transmit virus to fellow patients and unprotected health care workers ^1,2^. Moreover, undetected positive patients are more likely to develop perioperative pulmonary complications that are associated with a higher mortality after mayor surgery ^3^. Detection of COVID-19 positive cases is therefore crucial to protect patients and staff at the same time.

As an advice for the prevention of nosocomial transmission of COVID-19, the German Federal Institute for Public Health (Robert-Koch-Institut, RKI as of July 14th, 2020) provides the following guidance: “For inpatients without any recognizable complaints indicative of SARS-CoV-2 infection, testing according to a defined protocol before or during admission may be considered in order to minimize nosocomial transmissions. Decisions should take into account the prevailing epidemiological situation”^4^. However, it remains undecided how to adjust protocols to epidemiological contexts.

Currently, protocols for the screening of patients on hospital admission broadly fall into two categories, 1) algorithm-based screening (ABS), which pre-selects patients for further testing based on their symptoms and/or exposure history, and 2) universal admission screening (UAS), which tests all admitted patients to a certain hospital, department, or ward. Considerable differences exist in number of patients tested, costs and sensitivity, between these approaches, and it is not intuitive at what point in epidemiologically unstable situations it becomes advantageous to switch from one strategy to the other.

We therefore decided to develop a risk-adjusted decision tool, in the form of a nomogram, to inform hospitals with different admission rates when the introduction of an RT-PCR-based UAS on admission should be favoured over an ABS strategy. Crucially, our tool aims to take the evolution of background incidence in the regional population into account, offering a universal tool able to reconcile regional epidemiology with local hospital management decisions instead of focussing on nation-wide COVID-19 policies.

## Methods & Results

In order to make an informed decision between ABS and UAS, we need to estimate the sensitivity and specificity of both the preselection algorithm and the RT-PCR test, as well as the prevalence of COVID-19 in the study population.

We used data from the COVID-19 survey performed in the Italian town of Vó^5^ to determine the sensitivity and specificity of the pre-selection algorithm. This study surveyed almost all (85.9%) inhabitants of the Vó and tested them for SARS-CoV-2 using RT-PCR irrespective of their symptoms. Based on their first (cross-sectional) survey, 44 people had both symptoms and a positive PCR result, 149 had symptoms with a negative PCR results, 29 were asymptomatic PCR positive, and 2590 had no symptoms and a negative PCR result. Comparing the pre-selection algorithm to the RT-PCR results, the algorithm had a sensitivity of 60% and a specificity of 95%.

Estimates of the sensitivity of RT-PCR test are in the order of 70% to 80%^6,7^. In all likelihood, swabs occasionally fail to pick up viral RNA for different host-related or technical reasons. Specificity of the RT-PCR for SARS-CoV-2 viral RNA, on the other hand, is rather high (we here assume 99.99%) [ref]. However, not all PCR-positive patients produce infectious virus particles, and the specificity of RT-PCR for COVID-19 to identify active virus shedders i.e. infectiousness may therefore be considerably lower^8,9^.

We estimate the prevalence of infectious i.e. virus shedding, COVID-19-positive individuals (the primary goal of the screening strategies to avoid nosocomial spread) by multiplying the number of reported cases per 7 days in 100.000 population (weekly incidence) by a presumed average infectious period (8 days)^5,10,11^, divided by the estimated reporting proportion of 10%^12,13^. As a margin of safety, we purposely chose parameter estimates in a conservative manner such that the prevalence of infectious individuals in the community and underreporting is likely overestimated. The University Hospital Freiburg (UKF) has a catchment population of 658,419 inhabitants (the joined health districts of Freiburg, Breisgau-Hochschwarzwald, and Emmendingen), where 29 cases were reported during June 2020. This resulted in a mean prevalence of infectious individuals of 1.1 × 10^-4^.

The UKF admits about 73,000^14^ patients on an annual basis, on average 200 per day. Assuming a constant admission prevalence equal to the June community prevalence, the UKF would admit 8.3 infectious COVID-19 patients per year (In other words, on average one COVID-19 positive patient would be admitted every 44 days). ABS would detect 3.5 of these, and miss 4.8, while UAS would detect 5.8 and miss 2.5. At the same time, ABS would require testing of 3971 patients, while UAS would test all 73,000 patients.

To calculate the cost of improving the detection rates by switching from ABS to UAS, we calculated the Incremental Cost Effectiveness Ratio (ICER), defined as the additional cost (C_1_-C_0_) divided by the additional effect (E_1_-E_0_) of the new strategy (subscript 1), relative to the old strategy (subscript 0).

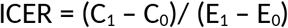

We define the cost of each strategy as the number of patients screened, and their effect as the number of true positive cases found. The ICER thus represents the number of additional patients screened to find one additional case, when switching from ABS to UAS.

Given a prevalence consistent with the regional epidemiology in June, 29983 patients need to be screened when switching to UAS to find a single additional case not yet identified by ABS. This involves screening of all patients admitted at UKF for nearly five months (149 days). At this prevalence, switching to UAS accomplishes only marginal risk reduction. As prevalence increase, however, thresholds for strategy changes start shifting.

To provide the means for hospitals to determine their risk of unwittingly admitting a patient not identified by ABS, the maximum number of days until that additional case would be identified by UAS can determined. This is a function of regional prevalence and the admission capacity of each hospital. First, we transform the ICER (R) by including the sensitivity of the algorithm (σ_a_) and the PCR test (σ_p_), the specificity of the algorithm (ρ_A_), the number of admissions per day (A), and the prevalence among admitted patients (P).

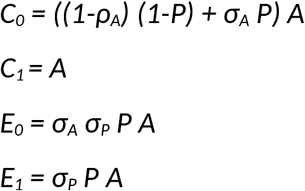

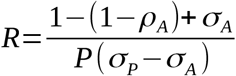

whereby, we can present the threshold as a function of the ICER:

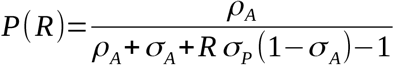

or as the number of days (D) screening all patients on admission:

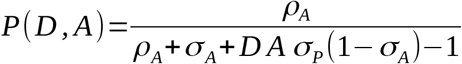

Using this function, a nomogram (figure) can be created. Each hospital can determine their own acceptable risk illustrated on the x-axes as the number of days until the first additional case would be detected by UAS but otherwise missed by ABS. Following the diagonal which corresponds to the hospital’s average daily admission rate until the intersection with their acceptable risk on the x-axes, the weekly incidence of regionally reported cases per 100,000 (on the y-axes) provides the threshold above which hospitals may want to switch from ABS to UAS.

**Figure 1:**
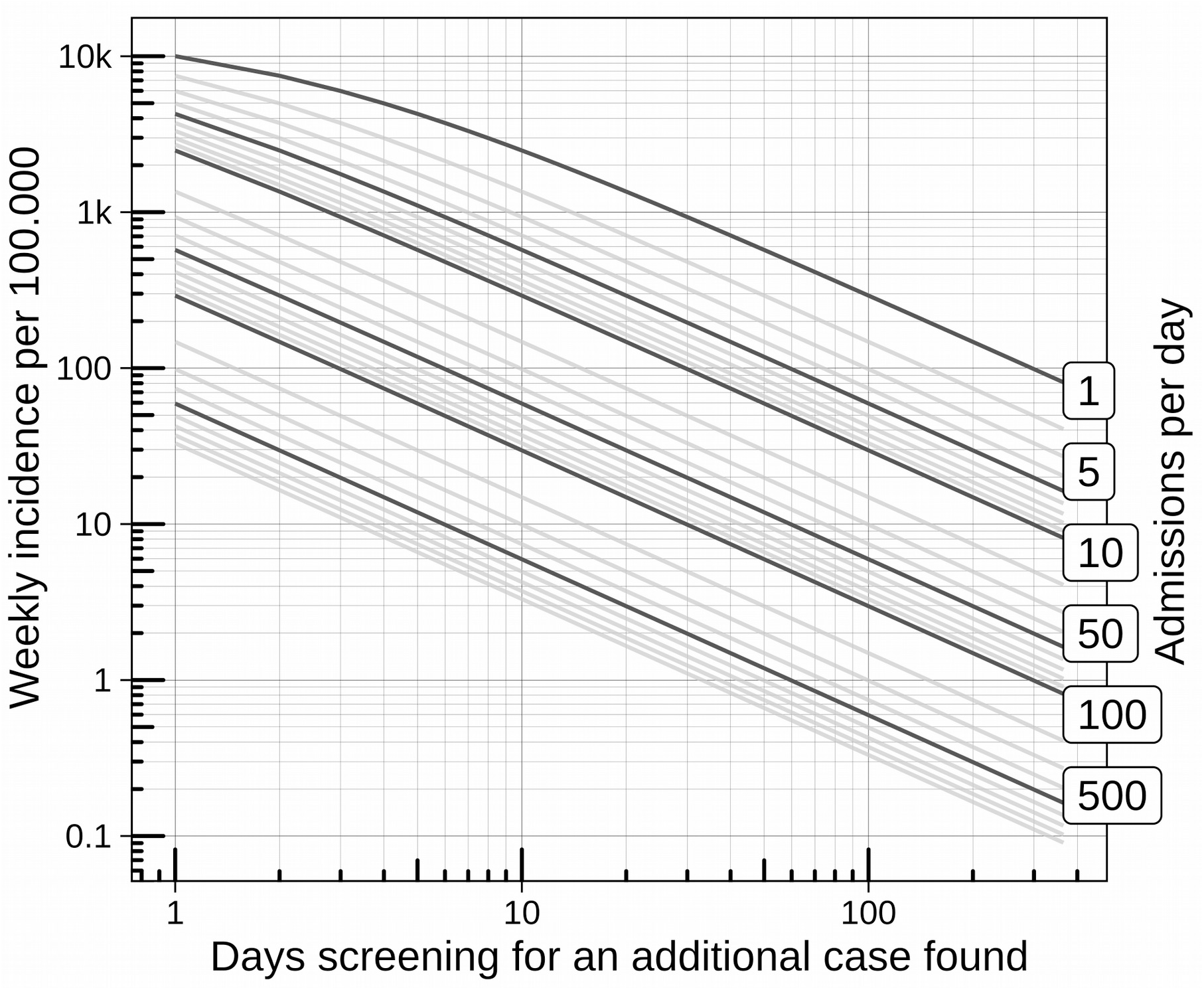
Nomogram to determine the weekly incidence threshold to switch from ABS to UAS, based on a hospital’s own acceptable risk illustrated on the x-axes as the number of days until the first additional case would be detected by UAS but otherwise missed by ABS. Following the diagonal, which corresponds to the hospital’s average daily admission rate, until the intersection with their acceptable risk on the x-aches the weekly incidence of regionally reported cases per 100,000 (on the y-axes) provides the threshold above which hospitals may want to switch from ABS to UAS.

## Discussion

In order to prevent nosocomial transmissions to patients and HCWs and to limit poor outcomes for undetected COVID-19 cases UAS for COVID-19 is widely advocated in hospitals serving high-incidence populations^1,15^. With declining or low incidence, however, the benefits of UAS are less clear. An alternative and more targeted approach consists of ABS. Thereby, only symptomatic patients or those with clearly defined risk factors are selected for testing. The question, however, arises, when to switch strategies against a background of unstable regional epidemiology.

In the absence of other quantitative appraisals of different strategies of testing for SARS-CoV-2, we here present a decision tool that takes into account three different variables, namely (i) the degree of risk to patients and HCW that can be averted, (ii) the pre-test probability or expected prevalence of infectious virus-shedding individuals in the hospital catchment population, and (iii) the average daily admission rate of the hospital. Potential benefits and harms must be carefully considered before accepting any level of risk for patients and HCW. Clearly, UAS has the benefit to inform HCWs in terms of PPE use, patient isolation or deferral of major non time-sensitive surgery. This benefit is diminished by the limited sensitivity of about 70-80% for singular RT-PCR test^6,7^. A potential harm of testing asymptomatic patients is the depletion of testing supplies and redirection of valuable hospital resources.

The nomogram (figure) offers a simple quantitative risk appraisal for considerations by hospital decision makers when to switch from algorithm-based screening to universal screening. By choosing conservative estimates for underreporting of COVID-19 cases in German communities and for the average period of SARS-CoV-2 communicability, our figure may overestimate the true risks.

## Data Availability

The presented analysis is based on publicly available data. All data sources are cited.

## Data availability

The presented analysis is based on publicly available data. All data sources are cited.

## References

1. Rickman HM, Rampling T, Shaw K, et al. Nosocomial transmission of COVID-19: a retrospective study of 66 hospital-acquired cases in a London teaching hospital. Clin Infect Dis. doi:10.1093/cid/ciaa816

2. Wu Z, McGoogan JM. Characteristics of and Important Lessons From the Coronavirus Disease 2019 (COVID-19) Outbreak in China: Summary of a Report of 72 314 Cases From the Chinese Center for Disease Control and Prevention. JAMA. 2020;323(13):1239-1242. doi:10.1001/jama.2020.2648

3. Mortality and pulmonary complications in patients undergoing surgery with perioperative SARS-CoV-2 infection: an international cohort study - The Lancet. Accessed August 7, 2020. https://www.thelancet.com/journals/lancet/article/PIIS0140-6736(20)31182-X/fulltext

4. Robert Koch Institut. Hinweise zur Testung von Patienten auf Infektion mit dem neuartigen Coronavirus SARS-CoV-2. https://www.rki.de/DE/Content/InfAZ/N/Neuartiges_Coronavirus/Vorl_Testung_nCoV.html

5. Lavezzo E, Franchin E, Ciavarella C, et al. Suppression of a SARS-CoV-2 outbreak in the Italian municipality of Vo’. Nature. Published online June 30, 2020:1-1. doi:10.1038/s41586-020-2488-1

6. Watson J, Whiting PF, Brush JE. Interpreting a covid-19 test result. BMJ. 2020;369. doi:10.1136/bmj.m1808

7. Wang W, Xu Y, Gao R, et al. Detection of SARS-CoV-2 in Different Types of Clinical Specimens. JAMA. 2020;323(18):1843-1844. doi:10.1001/jama.2020.3786

8. Ridgway JP, Shah NS, Robicsek AA. Prolonged shedding of severe acute respiratory coronavirus virus 2 (SARS-CoV-2) RNA among patients with coronavirus disease 2019 (COVID- 19). Infect Control Hosp Epidemiol. Published online undefined/ed:1-2. doi:10.1017/ice.2020.307

9. KCDC. Findings from investigation and analysis of re-positive case. KCDC. Accessed August 7, 2020. https://www.cdc.go.kr/board/board.es?mid=a30402000000&bid=0030

10. He X, Lau EHY, Wu P, et al. Temporal dynamics in viral shedding and transmissibility of COVID- 19. Nat Med. 2020;26(5):672-675. doi:10.1038/s41591-020-0869-5

11. Ashcroft P, Huisman JS, Lehtinen S, et al. COVID-19 infectivity profile correction. *ArXiv200706602 Q-Bio Stat.* Published online July 13, 2020. Accessed August 7, 2020. http://arxiv.org/abs/2007.06602

12. Stringhini S, Wisniak A, Piumatti G, et al. Repeated seroprevalence of anti-SARS-CoV-2 IgG antibodies in a population-based sample from Geneva, Switzerland. *medRxiv.* Published online May 6, 2020:2020.05.02.20088898. doi:10.1101/2020.05.02.20088898

13. RKI - Coronavirus SARS-CoV-2 - Serologische Untersuchungen von Blutspenden auf Antikörper gegen SARS-CoV-2 (SeBluCo-Studie). Accessed August 7, 2020. https://www.rki.de/DE/Content/InfAZ/N/Neuartiges_Coronavirus/Projekte_RKI/SeBluCo_Zwischenbericht.html

14. Zahlen und Fakten | Universitätsklinikum Freiburg. Accessed August 7, 2020. https://www.uniklinik-freiburg.de/uniklinikum/zahlen-und-fakten.html

15. Klompas M. Coronavirus Disease 2019 (COVID-19): Protecting Hospitals From the Invisible. Ann Intern Med. 2020;172(9):619-620. doi:10.7326/M20-0751

